# Efficiency of a Multi-step Screening and Enrichment Process for Prospective Sepsis Cohort Enrollment

**DOI:** 10.64898/2026.10.07.26364969

**Authors:** Lisa Cosimi, Wei Qi, Sarah Naz-McLean, Kastin Pan, Shilei Li, Hannah Laibinis, Alexis Rooney, Con Moran, Grace Valentine, Seungmin Hwang, Jacques Deguine, Joshua Pickard, Deborah Hung, Ann E. Woolley

## Abstract

**Background:** Host response and biomarker research to advance sepsis therapeutics requires well-characterized cohorts with longitudinal biospecimens across a range of hosts and infections. Automated screening based on SIRS criteria is sensitive but nonspecific and identifies more candidates than research teams can approach, sample, and follow longitudinally. We evaluated a structured enrichment review by non-physician study staff, done after automated screening in a hospital-wide prospective sepsis cohort.

**Methods:** The CISID cohort is a prospective observational study of hospitalized patients with suspected sepsis, enrolling adults from the ED, inpatient wards, and ICUs at a tertiary academic medical center. Patients were identified through: (1) automated electronic health record screening for two or more SIRS criteria with a 24-hour lookback, documentation of microbiologic culture or PCR, and receipt of a new antibiotic; (2) a sepsis enrichment step of structured chart review by trained non-physician study staff; and (3) discussion with the primary clinical team. Enrolled patients underwent physician adjudication into four diagnostic categories. To evaluate the enrichment step, we analyzed all patients screened during a 9-week period, assigned a composite “sepsis-likely” variable (CDC Adult Sepsis Event criteria or ICD-10 code R65.20/R65.21 within 48 hours before or after screening), and assessed mortality across each stage of screening. We also assessed concordance of the composite with physician adjudication among enrolled patients.

**Results:** From November 2023 through February 2026, 4,318 patients screened in through EHR-based automated screening; 1,220 (28%) were flagged for enrollment, and 288 were enrolled. Physician adjudication classified 209 (73%) as sepsis, 46 (16%) as infection without sepsis, 14 (5%) were classified as undifferentiated critical illness, and 19 (6.6%) as non-infectious sepsis mimics. Patients with sepsis were severely ill (median SOFA score, 9; ICU admission, 80%; shock, 48%), and 51% were immunocompromised; mortality was 14% in hospital and 27% at 90 days. In a 9-week sample of 907 screened encounters, 21% of those deferred and 70% of those flagged for enrollment met the sepsis-likely composite, a 3.3-fold increase in prevalence after structured study team review.

**Conclusions:** A brief structured enrichment review after automated screening tripled the prevalence of EHR-based sepsis indicators among patients selected for enrollment, while concentrating enrollment efforts on roughly one-quarter of screened-in patients in this hospital-wide, all-source suspected-sepsis cohort. This approach may help prospective cohorts direct limited biospecimen resources toward patients most informative for host-response and biomarker research.

## Background

Sepsis remains an enormous global burden with 49.9 million incident global cases reported in 2017.^1^ In the United States, sepsis accounts for ~1.7 million adult hospitalizations and at least 350,000 deaths annually.^2^ Despite improvements in diagnosis and clinical management in recent decades, hospital mortality has remained stable, exceeding 15% in the United States.^3^ Survivors of sepsis are at high risk for recurrent sepsis,^4,5^ readmission,^6–8^ and for longer-term cognitive and functional impairment.^9^

Sepsis is defined as a dysregulated host immune response to infection resulting in life-threatening organ dysfunction.^10^ This host response is now well-understood to be heterogeneous with evidence for distinct biological subgroups including transcriptomic endotypes, immune phenotypes and biomarker-defined states,^11–14^ as well as clinical phenotypes correlating with differing clinical outcomes and treatment responses.^15^ Together, this growing body of evidence supports the development of personalized targeted sepsis therapies. The Immunosep trial, which randomized patients with sepsis to therapy based on immune phenotype, and Tigris, which randomized those with septic shock and high endotoxin levels to Polymyxin B hemadsorption, demonstrated the potential benefit of precision therapies in practice, suggesting a future where optimized sepsis therapy will be individually tailored.^16,17^

Discovery of clinical and biological subgroups has been informed by numerous biospecimen-linked cohorts with varied approaches to screening and enrolling patients with suspected sepsis followed by downstream diagnostic confirmation. Notable examples include Intensive Care Unit (ICU) cohorts that enroll critically ill patients with suspected infection;^18,19^ and Emergency Department (ED) cohorts that employ clinical algorithms to identify patients with suspected sepsis upon presentation to the hospital. ^20,21^ Yield of confirmed or probable sepsis cases after downstream adjudication has generally reached approximately half in similarly designed cohorts.^18–20^ Other groups have enriched for sepsis cases by restricting enrollment to specific infections such as community-acquired pneumonia and fecal peritonitis.^12,22^

Further discovery and advancement toward a personalized therapeutic approach to sepsis is likely to require additional study and analyses of prospectively enrolled, well-characterized cohorts with linked longitudinal biospecimen sampling over a range of hosts, and over the spectrum and course of infection.^23^ Building such a diverse biospecimen cohort at scale requires balancing the sensitive but low-yield nature of screening processes triggered by the presence of signs of systemic inflammatory response syndrome (SIRS) plus suspected infection, against the finite resources required for informed consent, data collection and biospecimen processing.

Here we describe a hospital-wide, all-source suspected-sepsis cohort integrating a structured study team chart review, physician adjudication, longitudinal biospecimen linkage, and long-term clinical outcome measures in an immunocompromised-enriched tertiary hospital setting. The multi-step screening process efficiently enriches for individuals with sepsis, while the clinical adjudication links the spectrum of sepsis and non-sepsis categories to longitudinal biospecimens and clinical outcomes, at a breadth not previously described in earlier ICU, ED or infection-enriched cohorts.

## Methods

### Study design and setting

The Center for Integrated Solutions to Infectious Diseases (CISID) sepsis cohort is a prospective observational study of hospitalized patients with suspected sepsis designed to support host-response biologic discovery. The ongoing study is conducted at Brigham and Women’s Hospital, a tertiary academic medical center in Boston, Massachusetts, enrolling adults with suspected sepsis from the ED, inpatient wards, and ICUs. Enrollment began in November 2023 and is ongoing. This interim analysis includes patients enrolled through February 2026.

### Eligibility and screening

Eligible patients included adults (≥18 years) with suspected infection, defined as a new antibiotic administered, together with a microbiological culture or PCR obtained within the previous 24 hours and at least two SIRS criteria. Exclusions were limited to contraindications to biospecimen collection as determined by the study team or clinical team. Patients were identified through a three-layer screening process performed on weekdays that included (1) automated electronic medical-record screening, operationalized as daily automated electronic health record reports (Epic, Epic Systems Corporation, Verona, Wisconsin) spanning the ED and all inpatient locations with a 24-hour lookback for two or more signs of SIRS, microbiologic culture or PCR, and receipt of a new antibiotic; (2) a sepsis enrichment step whereby trained non-physician study staff performed structured chart review (estimated 3-5 minutes per chart) for evidence of positive microbiologic cultures or PCR results, chart documentation of confirmed or high clinical suspicion of sepsis by ICU or infectious disease consultants; and/or (3) in-person discussion with the primary clinical team. Individuals who passed all three steps were flagged for enrollment (Supplementary Table S1).

### Biospecimen collection

Whole blood was collected on up to three occasions, days 1, 3, and 7 (+/−1 day) post-enrollment, in patients who were alive and still admitted. Blood samples were processed the same day for peripheral blood mononuclear cells, plasma, and serum for downstream assays. Follow-up blood samples were collected at days 30 (+/−10), 90 (+/−21) and 180 (+/−28) from patients returning for clinical visits, or who remained in hospital, or were re-admitted during each respective window. Samples were also collected if a patient is admitted with any new occurrences of sepsis. All patients with at least one index sample in the first seven days were considered enrolled and retained for clinical-outcome analysis.

### Data collection and outcomes

Clinical data were abstracted from the electronic health record into REDCap electronic data capture tools hosted at Brigham and Women’s Hospital,^24,25^ and included demographics, comorbidities, medications, microbiology, and organ-dysfunction measures. Sequential Organ Failure Assessment (SOFA) scores were recorded at entry (defined as +/−12 hours from blood culture draw) and on days 1, 3, and 7. Outcomes included index hospitalization length of stay, ICU admission and ICU days, occurrence of shock, in-hospital and all-cause 30- and 90-day mortality, and 30- and 90-day readmission.

### Clinical adjudication

Enrolled patients underwent structured physician adjudication at least 14 days after enrollment to assess the clinical course, microbiological results, imaging, and treatment response. Two independent physician adjudicators applied a predefined rubric to determine the presence or absence of infection, the presence or absence of sepsis by Sepsis-3 criteria,^10^ the presence or absence of septic shock, and the anatomic and microbial source of infection. Discordant results were reviewed by a third physician tie-breaker.

Each patient was assigned to one of four categories: (i) adjudicated sepsis; (ii) infection not meeting Sepsis-3 criteria; (iii) undifferentiated critical illness (possible but unconfirmable infection e.g., cardiogenic presentation with possible aspiration pneumonia); (iv) non-infectious sepsis mimic (e.g. shock with alternative diagnosis and no infection identified).

### Screening and enrichment performance evaluation

In order to evaluate the performance of the manual screening step to enrich for patients with a higher likelihood for sepsis, we conducted a separate analysis of all patients screened for enrollment during a single 9-week period (7/15/2025-9/18/2025) to review the likelihood of sepsis diagnosis in each group of patients who were screened, screened out, screened-in, and flagged for enrollment after study team review. The review captured screening status and encounter-level data including blood cultures, antibiotics, organ dysfunction measures, International Classification of Diseases, Tenth Revision (ICD-10) codes, and mortality.

For each of the screened patients, we assigned a composite “sepsis-likely” variable which included presence of either an ICD-10 code for severe sepsis without or with septic shock (R65.20, R65.21) or (2) fulfillment of the Centers for Disease Control and Prevention (CDC) Adult Sepsis Event (ASE) criteria within 48 hours before or after screening.^26^ The CDC ASE is an automated, reproducible surveillance definition developed for hospital-level surveillance of sepsis that requires both evidence of presumed serious infection defined as a blood culture obtained and at least 4 days of antibiotics starting within two days before or after the blood draw, concurrently with the presence of acute organ dysfunction within the same window. The tool has a sensitivity of 69.7% (95% CI 52.9–92.0%) and a specificity of 98.1% (95% CI 97.7–98.5%) for predicting sepsis at a hospital-wide population level.^3,27^ Individuals in the 9-week screening enrichment analysis with either a positive ASE score or presence of a sepsis ICD-10 code (R65.20/R65.21) within 48 hours before or after screening were considered to be “sepsis-likely”. We assessed mortality and sepsis-likely composite positivity at each stage of screening.

### Statistical analysis

Cohort characteristics and outcomes are summarized descriptively by adjudicated diagnostic category as medians (IQR) and counts (percentages), with proportions reported with 95% Wilson confidence intervals. Statistical analysis was performed in R version 4.5.2. We also assessed concordance of the “sepsis-likely” composite with physician adjudication among enrolled patients and report positive predictive value, sensitivity, specificity with Wilson’s 95% confidence intervals.

### Ethics

The study was approved by the Mass General Brigham Institutional Review Board (Protocol 2023P000161), and all participants or their proxies provided written informed consent.

## Results

### Structured hospital-wide screening process yielded a cohort enriched for sepsis

Between November 6, 2023, and February 28, 2026, 4318 patients screened in by automated review for SIRS along with a new microbiologic culture and antibiotic in the previous 24 hours, of whom 1,220 (28%) met study criteria for suspected sepsis after structured chart review by the study team. Of these, 509 (42%) were approached, and 288 (57%) were consented, enrolled, adjudicated, and analyzed. Reasons for attrition included evidence of non-sepsis diagnosis upon study team review, care team or clinician declines, competing patient clinical priorities, competing study team priorities, lack of health care proxy, goals of care planning in progress, health care proxy or patient consent declines, and patient discharge or death prior to any biospecimen collection (Figure 1).

**Figure 1.**
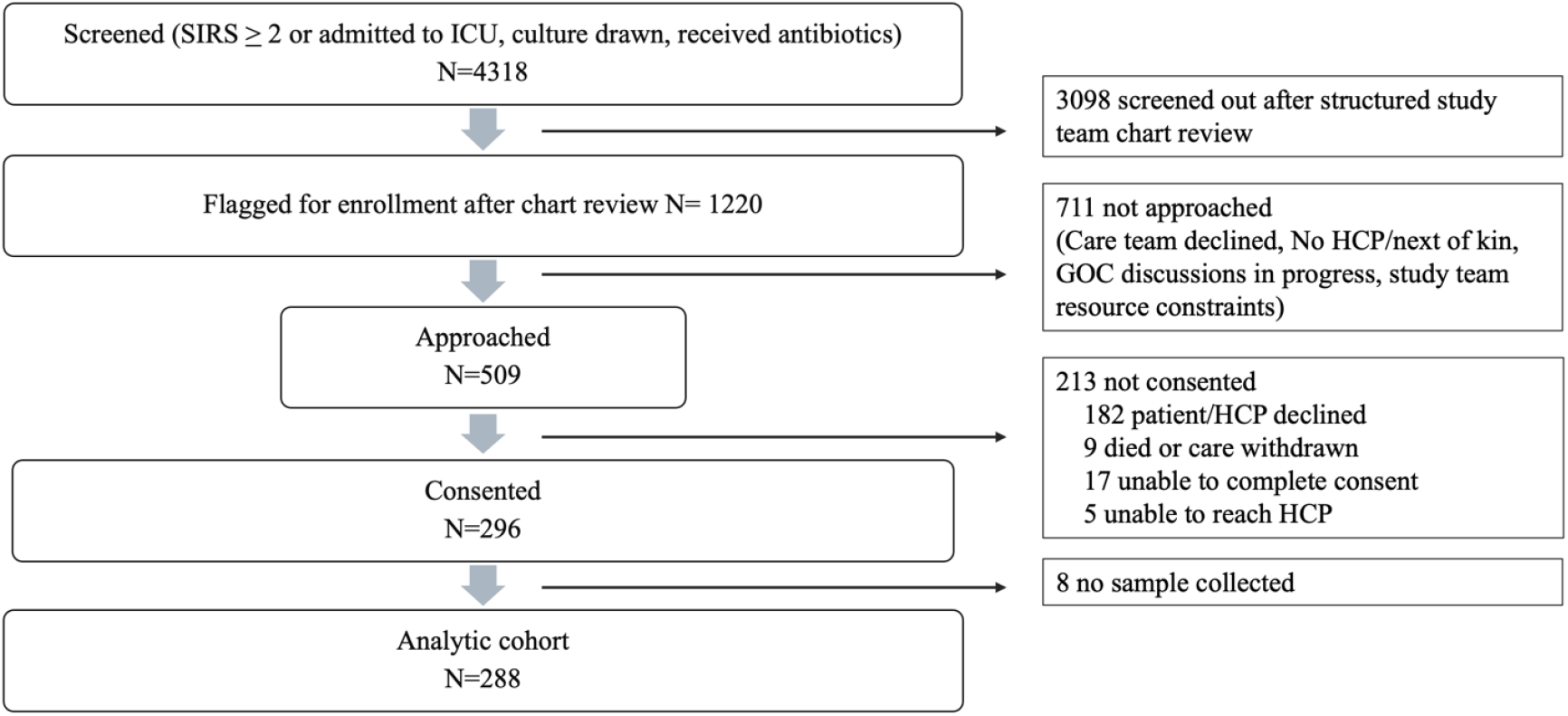
Screening and enrollment of the analytic cohort. Hospitalized patients were screened if they met at least two systemic inflammatory response syndrome (SIRS) criteria or were admitted to the ICU, had a culture drawn, and received antibiotics within the previous 24 hours (N = 4,318). The study team then conducted a structured chart review, which screened out 3,098 patients and flagged 1,220 for enrollment. Of the flagged patients, the team did not approach 711. Reasons included the care team declining, no available health care proxy or next of kin, ongoing goals-of-care discussions, and limited study team resources. Of the 509 patients approached, 213 did not consent: 182 patients or proxies declined, 9 died or had care withdrawn, 17 were unable to complete consent, and 5 had a proxy who could not be reached. Of the 296 patients who consented, 8 had no biospecimen collected, which left an analytic cohort of 288. GOC, goals of care; HCP, health care proxy; ICU, intensive care unit; SIRS, systemic inflammatory response syndrome.

Of the 288 adjudicated patients, 209 (73%) met Sepsis-3 criteria for sepsis, 46 (16%) were classified as having infection not meeting Sepsis-3 criteria, 14 (5%) had undifferentiated critical illness, and 19 (6.6%) were classified as non-infectious sepsis mimics.

### Clinical presentation, course, and outcomes of patients with sepsis

The median age of individuals with sepsis was 63 years (IQR 50-72), and 60% were male. Comorbidities included pulmonary disease in 101 (48%), chronic renal insufficiency in 75 (36%), diabetes in 78 (37%), and cardiac disease in 68 (33%) (Table 1).

**Table 1.** Characteristics of enrolled individuals overall and those with sepsis. BMI = Body Mass Index. * Not in remission. ** Based on CDC 2025 COVID-19 Vaccination Guidance for People Who Are Immunocompromised.^29^ *** Absolute neutrophil count < 500 cells/mm3 after chemotherapy; active hematologic malignancy, stem cell or solid organ transplant and on immunosuppression, prednisone equivalent > 20mg for > 2 weeks, receiving B cell inhibitor, bispecific antibody or anti-TNF inhibitor, HIV with CD4 < 200 cells/mm3, and primary immunodeficiency.

| **Characteristics** | **Total**<br>**N = 288**<br>**n (%)** | **Sepsis**<br>**N = 209**<br>**n (%)** |
| --- | --- | --- |
| **Demographics** |  |  |
| Age (years) - median (IQR) | 63 (48, 72) | 63 (50, 72) |
| BMI (kg/m<sup>2</sup>) - median (IQR) | 27 (23, 31) | 27 (23, 31) |
| Sex assigned at birth |  |  |
| Male | 168 (58) | 126 (60) |
| Female | 120 (42) | 83 (40) |
| Race/Ethnicity |  |  |
| White Non-Hispanic | 207 (72) | 151 (72) |
| Black Non-Hispanic | 21 (7) | 19 (9) |
| Hispanic or Latinx | 28 (10) | 19 (9) |
| Asian Non-Hispanic | 11 (4) | 8 (4) |
| Other | 3 (1) | 2 (1) |
| Unknown | 18 (6) | 10 (5) |
| **Comorbidities** |  |  |
| Cardiac disease | 83 (29) | 68 (33) |
| Hypertension | 180 (63) | 136 (65) |
| Diabetes | 100 (35) | 78 (37) |
| Stroke | 37 (13) | 30 (14) |
| Pulmonary disease | 139 (48) | 101 (48) |
| Autoimmune disease | 51 (18) | 38 (18) |
| Cirrhosis | 20 (7) | 14 (7) |
| Chronic renal insufficiency | 97 (34) | 75 (36) |
| On dialysis | 19 (7) | 16 (8) |
| History of malignancy | 175 (61) | 124 (59) |
| Solid tumor* | 77 (27) | 53 (25) |
| Hematologic malignancy* | 50 (17) | 34 (16) |
| Stem cell transplant | 22 (8) | 17 (8) |
| Solid organ transplant | 29 (10) | 18 (9) |
| Immunocompromised** | 152 (53) | 107 (51) |
| Moderate to severe immunocompromise*** | 98 (34) | 64 (31) |
| **Pre-Hospitalization Exposures** |  |  |
| Recent surgery <30 days | 69 (24) | 50 (24) |
| Hospitalized in last 30 days | 81 (28) | 56 (27) |
| Sepsis related | 13 (5) | 10 (5) |
| Hospitalized in last 180 days | 187 (65) | 139 (67) |
| Sepsis related | 47 (16) | 38 (18) |

Over half of the adjudicated sepsis cohort (51%) was immunocompromised, including 64 (31%) who were moderate-to-severely immunocompromised (defined as having an absolute neutrophil count < 500 cells/mm3 after chemotherapy, active hematologic malignancy, stem cell or solid organ transplant and on immunosuppression, prednisone daily equivalent > 20mg for > 2 weeks, receiving B cell inhibitor, bispecific antibody or anti-TNF inhibitor, HIV with CD4 < 200 cells/mm3, or primary immunodeficiency).^28^ Healthcare exposures were common in the 30 days prior to the index hospital admission, including 56 (27%) hospitalizations, of whom 22 (11%) were admitted with infection and 10 (5%) with sepsis, and 50 (24%) who had undergone a surgical procedure. In the 180 days prior to enrollment, 139 (67%) had been hospitalized (Table 1).

The most common anatomic source of infection was pulmonary (31%) followed by urinary (15%), hepatobiliary (15%), gastrointestinal (14%), vascular including endocarditis and central line associated infections (13%), followed by bone or joint (3%), central nervous system (1%) and genital (1%). A microbial source of infection was confirmed in 144 (69%) individuals. Gram-negative infections occurred in 80 (38%), followed by Gram-positive infections in 73 (35%), and anaerobes in 12 (6%). Viral and fungal sources were present in 11 (5%) and 7 (3%), respectively, and 42 (20%) presented with polymicrobial infections. The most common pathogens included *Escherichia coli, Klebsiella spp*., *Pseudomonas aeruginosa, Staphylococcus aureus, Enterococcus spp*., *Enterobacter spp*., and *Candida spp*. (Table 2).

**Table 2.**
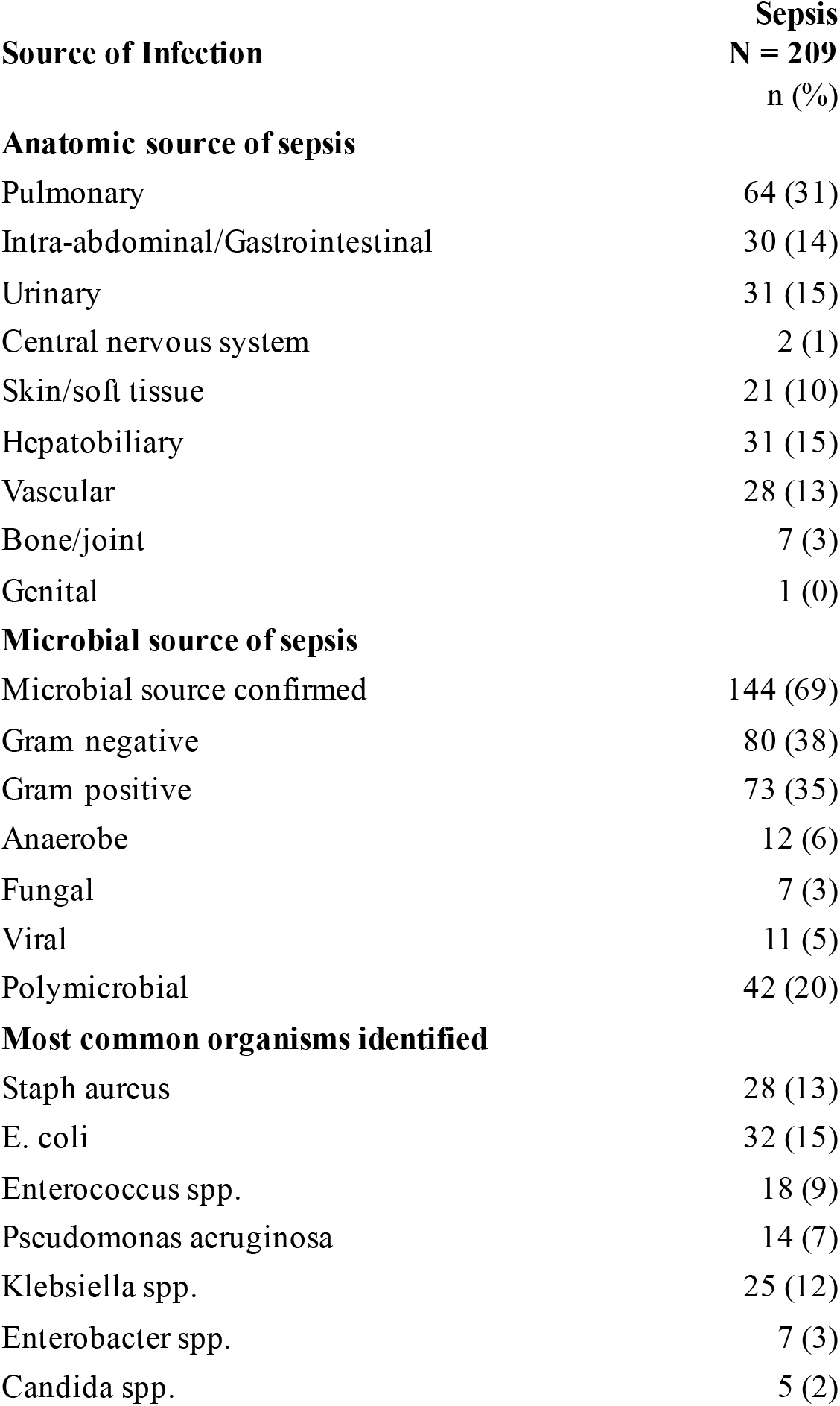
Anatomic and microbial sources of infection in individuals adjudicated with sepsis based on Sepsis-3 criteria.

Most individuals adjudicated with sepsis were enrolled from the intensive care unit (71%), while 17% and 12% entered from the emergency department and inpatient floors, respectively. Severity of illness within the sepsis group was high, with a median entry Sequential Organ Failure Assessment (SOFA) score of 9 (IQR, 7-12), 168 (80%) requiring intensive care unit (ICU) admission and 100 (48%) with shock. The median hospital length of stay was 14 days (IQR, 7-28). In-hospital mortality was 14% and rose to 17% at 30 days and 27% at 90 days.

Among the 179 patients who survived the index hospitalization, 66 (37%) were readmitted within 30 days. Of these, 33 (18%) were admitted with infection and 18 (10%) with sepsis. Readmission rates rose to 53% by 90 days, with 15% readmitted with sepsis. Of patients readmitted, the median time to readmission was 21 days (IQR, 11-66) (Table 3).

**Table 3.** Clinical severity and outcomes in enrolled individuals adjudicated with sepsis.

| **Clinical Severity and Outcomes** | **Sepsis**<br>**N = 209**<br>**n (%)** |
| --- | --- |
| **Status at entry** |  |
| Floor | 26 (12) |
| ICU | 148 (71) |
| ED | 35 (17) |
| Entry SOFA score | 9 (7, 12) |
| **Hospital outcomes** |  |
| Length of stay (Median, IQR) | 14 (7, 28) |
| Admitted to ICU | 168 (80) |
| Days in ICU (Median, IQR) | 3 (1, 9) |
| Shock | 100 (48) |
| **Mortality** |  |
| In-hospital | 30 (14) |
| 30-day | 35 (17) |
| 90-day | 57 (27) |
| **Readmission (survivors)** | **(n = 179)** |
| 7-day | 16 (9) |
| 30-day | 66 (37) |
| with sepsis | 18 (10) |
| 90-day | 95 (53) |
| with sepsis | 26 (15) |
| Days until readmission (Median, IQR) | 21 (11, 66) |

### Enrichment Analysis Cohort

#### Screening and structured study team review enriched for individuals with sepsis and remained representative of the screened population

Of 907 screened patients assessed to evaluate the effectiveness of the structured enrichment process over a single 9-week period, 500 (55%) were screened out due to lack of either antibiotic administration or microbiologic culture in the previous 24 hours, 303 (33%) screened in, but were deferred after study team review, and 104 (11%) were flagged for enrollment (Figure 2). The sepsis-likely composite (CDC Adult Sepsis Event criteria or ICD-10-CM code R65.20/R65.21 within ±48 hours of screening) was met in 55 of 500 (11%) screened-out patients, 64 of 303 (21%) deferred patients, and 73 of 104 (70%) patients flagged for enrollment, a 3.3-fold increase in flagged vs. deferred patients.

**Figure 2.**
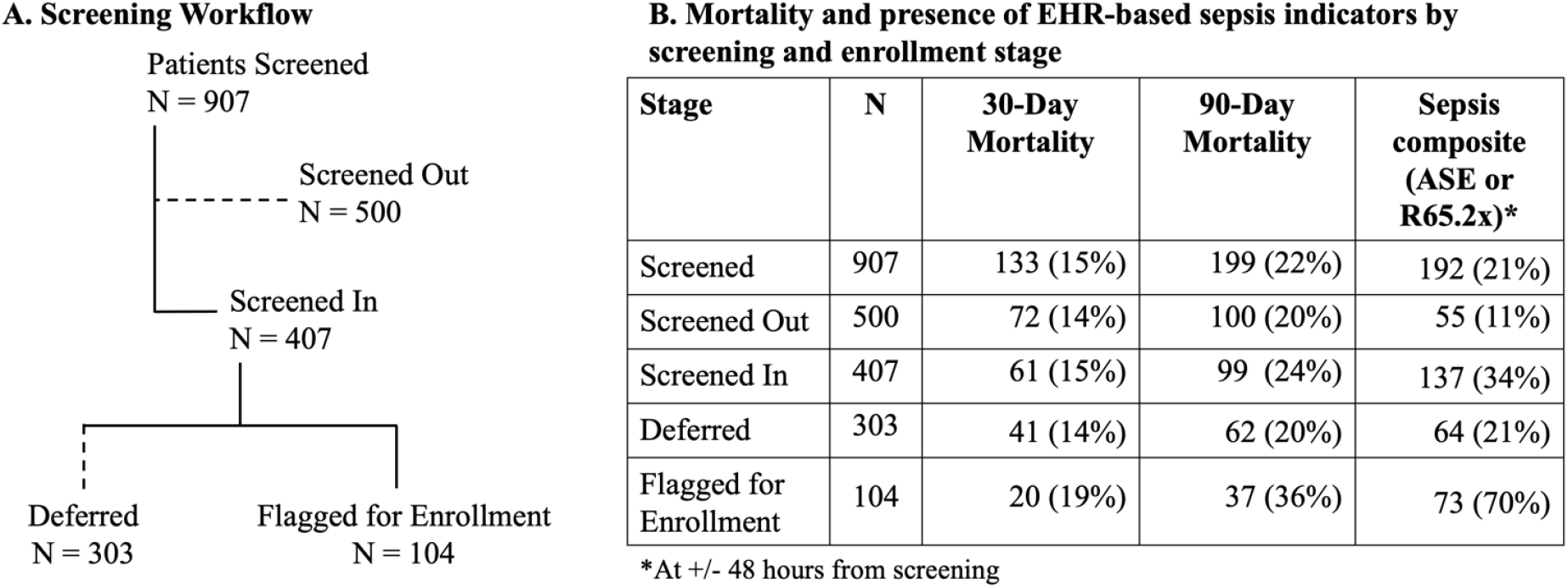
Mortality and sepsis outcomes by screening and enrollment stage among patients screened for sepsis at Brigham and Women’s Hospital Patients screened for sepsis in a 9-week enrichment analysis (N = 907) were classified as screened out (n = 500) or screened in (n = 407). Screened-in patients were further classified as deferred (n = 303) or flagged for enrollment (n = 104) after structured study team review. Percentages use the row N as the denominator. Mortality is all-cause death within 30 and 90 days of screening. The composite variable is the presence of a CDC Adult Sepsis Event (ASE) or an ICD-10-CM code for severe sepsis (R65.20) or septic shock (R65.21) within ±48 hours of screening. ASE, Adult Sepsis Event; ICD-10-CM, International Classification of Diseases, Tenth Revision, Clinical Modification.

There was minimal difference in 30-day mortality across the screening stages, with 72/500 (14%), 41/303 (14%), and 20/104 (19%) in the screened out, deferred, and flagged for enrollment groups respectively. However, 90-day mortality was higher, among flagged patients (37/104, 36%), compared with 62/303 (20%) in the deferred group and 100/500 (20%) in those screened out.

We next compared the sepsis-likely composite to physician adjudication among the 288 enrolled patients from the analytic cohort enrolled November 2023 through February 2026. Of these, 190 (66%) also met the sepsis-likely composite. Relative to physician adjudication, the composite had a positive predictive value of 87% (165/190; 95% CI, 81%–91%), sensitivity of 79% (165/209; 95% CI, 73%–84%), and specificity of 68% (54/79; 95% CI, 58%–78%) (Supplementary Table S2). The 25 composite-positive patients not adjudicated as sepsis included 10 (40%) with infection not meeting Sepsis-3 criteria, 10 (40%) with undifferentiated critical illness, and 5 (20%) non-infectious mimics.

## Discussion

The CISID sepsis cohort is an ongoing, hospital-wide, all-source, prospective study of individuals with suspected sepsis incorporating a structured chart review step designed to enrich for patients with sepsis. The manual enrichment step captured clinical documentation, working diagnoses, and clinical trajectory, which EHR screens did not capture well.

Prospective sepsis cohorts face practical challenges: EHR-based screening built on SIRS criteria, cultures, and antibiotic initiation is sensitive but nonspecific,^10^ generating far more screen-ins than research staff can approach, consent, and sample. Without a second step, enrollment capacity may be spent largely on patients later found not to have sepsis. In a 9-week sample of 907 screened encounters, the structured screening step resulted in a 3.3-fold increase in the prevalence of the sepsis-likely composite. The step thus enriched the approached population toward patients with a higher pretest probability of sepsis, focusing consent, biospecimen collection, processing, data management, and longitudinal follow-up efforts on the cases most likely to be informative for the cohort’s specific aims. Because the 9-week sample captured only a portion of the enrollment period, we compared it with the full study. The proportion of screened-in patients flagged at review was similar in the 9-week sample (26%; 104/407) and across the full enrollment period (28%; 1,220/4,318), and 73% of enrolled patients were adjudicated as having sepsis. Together, these findings suggest the enrichment observed in the 9-week sample reflects how the process performs throughout our study.

The enrichment analysis highlighted a potential trade-off of the multistep screening process: 119 of 192 composite-positive individuals in the 9-week enrichment analysis cohort were not flagged for enrollment; 55 screened out at the electronic screening step, and 64 were deferred at chart review. However, composite positivity likely overstates the number of individuals with sepsis. The CDC Adult Sepsis Event tool was developed and validated as a surveillance definition for estimating sepsis incidence at the population level across hospitals where it performs well against clinical review.^3,27^ Here we combined it with ICD-10 codes for severe sepsis and septic shock for a different purpose: as a proxy within a population already selected for suspected infection where empiric antibiotics are routine and organ dysfunction may have non-infectious causes. Consistent with this, roughly one in three enrolled patients not adjudicated sepsis met the composite, so many of the unflagged composite-positive patients likely did not have sepsis. The review step prioritized enrolling patients most likely to have sepsis over enrolling every case. The enrolled sepsis cohort reflects this design. Patients were severely ill, most requiring ICU admission, nearly half with shock and with a mortality of 27% at 90 days, and readmission common among hospital survivors.

Unlike ICU-based cohorts that enroll consecutively and adjudicate afterward,^18^ or enrichment designs that select on immune phenotype,^16^ our approach enriches on clinical likelihood of sepsis before enrollment across ED, ward, and ICU settings, retaining physician adjudication as the final diagnostic step. To our knowledge, few prospective cohorts have reported yield at each step of their screening process; doing so may help investigators weigh enrichment against completeness when designing enrollment strategies.

## Limitations

We acknowledge several limitations. First, the screening enrichment analysis relied on EHR-based indicators rather than physician adjudication, which was not feasible for every encounter. Composite positivity among individuals not flagged for enrollment should therefore be interpreted as a surveillance-based proxy rather than missed diagnoses. Conversely, given the composite’s imperfect sensitivity, some sepsis cases may have occurred among individuals who were not flagged for enrollment. Second, the 9-week enrichment analysis sample and the enrolled cohorts came from different, though overlapping periods. Comparisons between the groups are therefore indirect, because the patients largely differ across time periods. Third, the structured enrichment review required initial training of study staff and dedicated staff time (3-5 minutes per chart) for each review. These costs are likely offset by the enrollment effort they save, because consent, biospecimen collection, and follow-up are concentrated on fewer, higher-yield patients, but we did not formally measure this. Performance and time may also vary with staff experience and between staff. Finally, this was a single-center analysis in a tertiary hospital with a high burden of immunocompromise, which may limit generalizability.

## Conclusion

In this prospective, hospital-wide sepsis cohort, a brief structured enrichment review by trained non-physician study staff after SIRS-based electronic screening identified patients with more than threefold higher prevalence of EHR-based sepsis indicators than those deferred, while concentrating enrollment efforts on roughly one-quarter of screened-in patients. For prospective cohorts, which may be limited by resources for consent, sample collection, data management, and longitudinal follow-up, a similar review step may help ensure that resources are directed toward the patients most informative for host-response and biomarker research.

## Data Availability

All data produced in the present work are contained in the manuscript

