## Supplementary Table 1 for "Efficiency of a Multi-step Screening and Enrichment Process for Prospective Sepsis Cohort Enrollment"

**Supplementary Table S1 EHR reports**

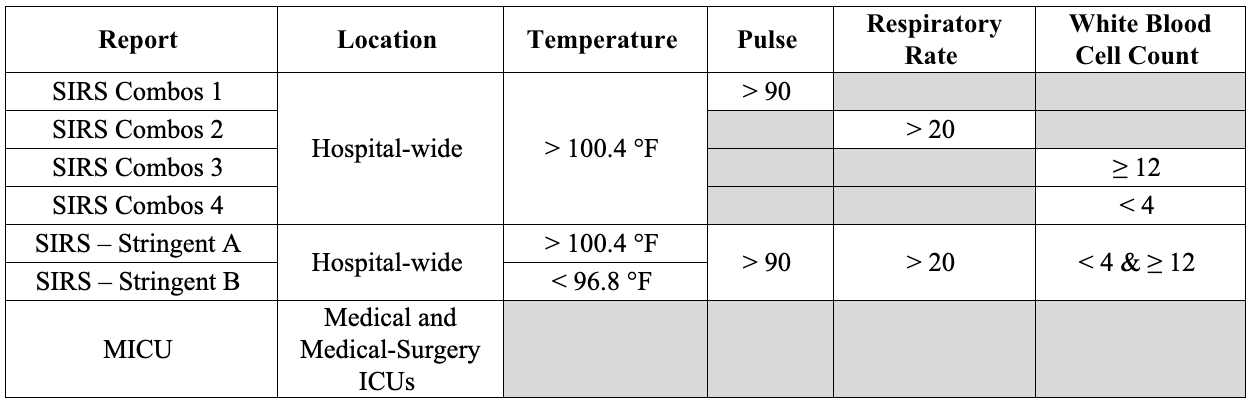
**Supplementary Table S1** Automated EHR reports used to screen patients for cohort eligibility. All patients resulting from the reports are at least 18 years old, and may be admitted to the hospital floors, ICUs, or emergency department.
