## Supplementary Table 2 for "Efficiency of a Multi-step Screening and Enrichment Process for Prospective Sepsis Cohort Enrollment"

**Supplementary Table S2**

| **A. Sepsis-likely composite present in individuals adjudicated sepsis or not sepsis** | | | |
| --- | --- | --- | --- |
|  | **Adjudicated sepsis** | **Not sepsis** | **Total** |
| Composite positive | 165 | 25* | 190 |
| Composite negative | 44 | 54 | 98 |
| Total | 209 | 79 | 288 |
| **B. Test statistics of sepsis-likely composite compared to physician adjudication** | | | |
|  | **Value** | **95% CI (Wilson)** | |
| Positive predictive value | 165/190 (87%) | 81%-91% | |
| Sensitivity | 165/209 (79%) | 73%-84% | |
| Specificity | 54/79 (68%) | 58%-78% | |
| Negative predictive value | 54/98 (55%) | 45%-65% | |

**Supplementary Table S2. Performance of the EHR-based sepsis-likely composite compared with physician-adjudicated sepsis.**

The sepsis-likely composite was defined as a CDC Adult Sepsis Event or an ICD-10-CM code for severe sepsis (R65.20) or septic shock (R65.21) within ±48 hours of screening. We compared it against physician adjudication of sepsis by Sepsis-3 criteria in the enrolled analytic cohort (N = 288). (A) Cross-tabulation of composite status against adjudicated sepsis status. (B) Positive predictive value, sensitivity, specificity, and negative predictive value of the sepsis-likely composite, with 95% confidence intervals calculated by the Wilson score method. Adjudicated sepsis prevalence in the enrolled cohort was 73% (209/288). Among the 25 sepsis-likely composite-positive patients not adjudicated as having sepsis, 10 (40%) had infection that did not meet Sepsis-3 criteria, 10 (40%) had undifferentiated critical illness, and 5 (20%) were non-infectious mimics. ASE, Adult Sepsis Event; CI, confidence interval; EHR, electronic health record; ICD-10-CM, International Classification of Diseases, Tenth Revision, Clinical Modification.
